# Study Protocol: Prognostic Factors of High Healthcare Utilization Costs Among People with Spinal Disorders Using a Spine Registry Linked to Public Healthcare Data

**DOI:** 10.1101/2025.08.05.25333009

**Authors:** Daniel Feller, Bjørnar Berg, Ørjan Nesse Vigdal, Lise Grethe Kjønø, Maja Wilhelmsen, Bart Koes, Margreth Grotle, Alessandro Chiarotto

## Abstract

**Objective:** To identify prognostic factors associated with high spine-related healthcare costs among patients referred to secondary care for spinal disorders in Norway.

**Study Design and Setting:** This prognostic factor study will follow the PROGnosis RESearch Strategy (PROGRESS) framework. We will utilize data from 7,877 patients included in the Norwegian Neck and Back Registry between 2016 and 2020, linked to national administrative databases, to estimate spine-related healthcare costs over a 12-month period following the index consultation.

**Methods:** Candidate prognostic factors will include demographic, clinical, psychosocial, and work-related variables collected at baseline. Healthcare costs will be estimated using reimbursement rates from national registries and adjusted to 2022 price levels. Missing data will be handled through multiple imputation using chained equations. Associations between predictors and costs will be assessed using generalized linear models with a Gamma distribution. Continuous variables will be modeled using restricted cubic splines. All models will be adjusted for prior healthcare costs, age, gender, baseline pain intensity, disability, and other significant univariable predictors. Subgroup analyses (e.g., by pain location) and sensitivity analyses (complete-case analysis) will be conducted.

**Conclusion:** This study will provide evidence on factors associated with high spine-related healthcare costs, aiming to inform future research and health policy.

## INTRODUCTION

Spinal disorders are among the leading causes of disability worldwide, representing a significant public health concern across countries and healthcare systems. ^1–3^ According to the 2021 Global Burden of Disease Study, low back pain (LBP) consistently ranks as the most disabling condition globally, with neck pain also among the top 10 contributors to Years Lived with Disability. ^1,2,4^ These conditions impose a substantial burden not only in terms of individual suffering but also in economic terms, including costs associated with healthcare utilization. ^5–8^ In fact, spinal disorders are associated with significant healthcare expenditures, related to : primary care consultations, pharmacological treatments, diagnostic imaging, specialist referrals, and hospital admissions. ^9,10^ Notably, many of these healthcare services are not aligned with current clinical practice guidelines. ^11,12^ For example, imaging for acute LBP is frequently prescribed despite strong recommendations against its routine use in the absence of red flags, highlighting a persistent issue of overuse/overdiagnosis and low-value care. ^13,14^ It is important to highlight that the existing literature suggests that the majority of healthcare costs associated with spinal disorders are generated by a relatively small subset of patients. ^15^

This high-cost subgroup often utilizes disproportionate amounts of healthcare resources, yet the factors predicting such utilization patterns remain insufficiently understood. Identifying prognostic factors for high-cost healthcare users among patients with spinal disorders could offer valuable insights for designing targeted interventions for cost reduction and more efficient allocation of healthcare resources. Previous studies have explored prognostic factors associated with high healthcare utilization in individuals with musculoskeletal conditions. For instance, Killingmo et al. ^10^ investigated modifiable predictors of high healthcare costs among older adults consulting primary care for back pain, while Mutubuki et al. ^16^ examined factors associated with high societal costs in patients with chronic LBP. However, the available evidence remains limited in terms of generalizability and does not fully capture the complexity of cost drivers across the spectrum of spinal disorders. In this context, we planned a prognostic factor study to investigate the variables associated with high healthcare utilization costs among patients with spinal disorders. ^17^ By identifying predictors of high healthcare costs, this study aims to inform clinical decision-making and health policy strategies, ultimately contributing to the provision of high-value, cost-effective care for individuals with spinal disorders.

## METHODS

This study is part of the AID-Spine project, which has been approved by the ethics committee of the Health Region of South-East Norway (2022/371282), and the Data Protection Authority of Norway approved the registry protocol. All participants gave written informed consent upon participation in the NNRR, including consent to link their data to other registries.

This study is designed and conducted in accordance with the “PROGnosis RESearch Strategy” (PROGRESS) framework, with particular reference to Part 2, which focuses on prognostic factor research. ^17^ The reporting of the manuscript will be guided by the “REporting recommendations for tumor MARKer prognostic studies” (REMARK) checklist, adapting its principles to the context of non-oncological prognostic research. ^18^ The nature of this study is exploratory, and no a priori hypotheses will be made regarding the associations between potential prognostic factors and the outcome. ^19^

### Data sources

This study will leverage data from the Norwegian Neck and Back Registry (NNRR), a large, national clinical registry designed to systematically collect data on patients referred to a specialist assessment in secondary care receiving conservative management due to spinal disorders (i.e., back and/or neck pain). ^20^ The NNRR was established in 2014 as a national medical quality registry to address the growing need for systematic data on patients with neck and back complaints assessed in specialist outpatient care. ^20^ As of 2023, the registry includes data from 14 multidisciplinary outpatient clinics across Norway. ^20^ These clinics are integrated within hospital-based outpatient physical medicine and rehabilitation departments and are mandated to deliver specialist care involving interdisciplinary teams. Patients are predominantly referred to the outpatient clinics by their general practitioner, although referrals may also originate from manual therapists, other specialist physicians, or psychologists. Upon receiving a referral, a senior physician assesses patient eligibility based on standardized national criteria. ^20^ To qualify for inclusion in the registry, patients must be 18 years or older and present with a primary complaint related to spinal pain. ^20^ Patients fill out a digital questionnaire just prior to the first consultation and at 6 and 12 month follow-up. After their initial clinic visit, the attending clinician completes a standardized report detailing the consultation. Before 2020, follow-up was conducted by mail at 6 months. ^20^ Up to 2023, the registry has accumulated data from 18,982 consenting patients. ^20^ Among these, 12,741 reported neck pain, 11,510 reported thoracic pain, and 15,397 reported LBP. ^20^ For this project, we will use data collected between January 1st, 2016, and December 31st, 2020, comprising a total of 9,354 patients (63% with back pain, 27% with neck pain, and 10% with both back and neck pain). From this cohort, we will exclude patients without a spine-specific International Classification of Diseases (ICD-10) code (n = 1,371) (Appendix 1). We will exclude patients for whom the index consultation could not be identified in the healthcare registry data (n = 87). Those who emigrated or died during follow-up will also be excluded based on data from Statistics Norway (n = 19). Therefore, the total sample size for our study will be 7,877 (Figure 1).

Table 1 reports variables collected in the NNRR on: patient’s characteristics at first consultation, employment characteristics at first consultation, general health at first consultation, treatment characteristics, and other musculoskeletal complaints at first consultation.

**TABLE 1.** VARIABLES COLLECTED IN THE NNRR DATASET.

|  |
| --- |
| **TABLE 1 – VARIABLES COLLECTED IN THE NNRR DATASET** |
| Age (in years) |
| Gender |
| *Male* |
| *Female* |
| Educational level among 25–66 years old |
| *Primary school, 7–10 years* |
| *Vocational school* |
| *Senior high school* |
| *College or university (<4 years)* |
| *College or university ($\geq 4$ years)* |
| Marital status |
| *Married* |
| *In a relationship* |
| *Single* |
| Number of own children |
| *0* |
| *1* |
| *2* |
| *3 or more* |
| Number of children in household |
| *0* |
| *1* |
| *2* |
| *3 or more* |
| Nationality |
| *Norwegian* |
| *Non-Norwegian European* |
| *Other* |
| Reading and writing difficulties |
| *Yes* |
| *No* |
| Regional health authority |
| *South-East Norway* |
| *Western Norway* |
| *Central Norway* |
| *Northern Norway* |
| *Unknown* |
| Currently working full time |
| *Yes* |
| *No* |
| Statuses |
| *Working* |
| *On sick leave* |
| *Sick leave 100%* |
| *Average sick leave* |
| *Work assessment allowance* |
| *Disability benefit* |
| *Disability benefit 100%* |
| *Student* |
| *Unemployed* |
| *Homemaker (unpaid)* |
| Applied for disability benefits |
| *Yes* |
| *No* |
| Applied for work assessment allowance |
| *Yes* |
| *No* |
| Sick leave same condition past 2 years |
| *Yes* |
| *No* |
| Perception that employer would like patient back at work |
| *Yes* |
| *No* |
| Self-reported work capability (scale 0–10) |
| Physical job demands (scale 1 – 5) |
| Mental job demands (scale 1 – 5) |
| Job satisfaction (scale 0–10) |
| Physical activity |
| *Physically inactive* |
| *Light physical activity* |
| *Regular physical activity and training* |
| *Hard physical training* |
| Smoking daily |
| *Yes* |
| *No* |
| Over-the-counter painkillers daily |
| *Yes* |
| *No* |
| Prescription painkillers daily |
| *Yes* |
| *No* |
| Hopkins symptoms checklist (HSCL-1) |
| Subjective Health Complaints |
| *Headache* |
| *Neck pain* |
| *Upper back pain* |
| *Lower back pain* |
| *Shoulder pain* |
| *Sleep problems* |
| *Tiredness* |
| Health today (visual analogue scale) |
| EQ5D-3L or EQ5D-5L |
| Initial diagnostic triage: neck |
| *Non-specific conditions* |
| *Neurological dysfunctions* |
| *Specific diagnosis for neck pain* |
| Initial diagnostic triage: back |
| *Non-specific conditions* |
| *Neurological dysfunctions* |
| *Other specific diagnoses for back pain* |
| *Additional diagnosis* |
| First consultation conducted by |
| *Doctor* |
| *Nurse* |
| *Physical therapist* |
| *Other clinical profession* |
| Patient use of opioids, antidepressants, sedatives, GABA medications, paracetamol, or non-steroidal anti-inflammatory drugs |
| *Yes* |
| *No* |
| Duration of current complaints |
| *No pain* |
| *Less than 3 months* |
| *3–12 months* |
| *1–2 years* |
| *>2 years* |
| Perceived causes of pain |
| *Workload* |
| *Load at home* |
| *Emotional distress* |
| *Leisure activities* |
| *Skeletal injury* |
| *Muscular injury* |
| *Nerve injury* |
| *Malpractice* |
| *Don't know* |
| Oswestry Disability Index (0%–100) |
| Neck Disability Index (0%–100) |
| Fear-Avoidance Beliefs for Back Pain (Physical Activity and Work subscales) |
| Location of painful body regions |
| *Head* |
| *Neck* |
| *Lower back* |
| Pain at rest (0 – 10) |
| Pain during activity (0 – 10) |

Cost data will be obtained by linking the NNRR dataset to three national administrative databases: the Norwegian Control and Payment of Health Reimbursements Database, the Norwegian Patient Registry, and the Norwegian Prescription Database. ^21^ The Norwegian Control and Payment of Health Reimbursements Database contains information on reimbursement claims from general practitioners, physiotherapists, chiropractors, and emergency care clinicians in primary care settings, covering both face-to-face visits and teleconsultations. ^21^ The Norwegian Patient Registry provides data on publicly funded specialist healthcare services, including those delivered by private institutions and medical specialists under contract with the regional health authorities. ^21^ The Norwegian Prescription Database is a nationwide registry that contains information on all prescription drugs dispensed at pharmacies to individuals in Norway. This database includes data on the type of medication (classified according to the Anatomical Therapeutic Chemical system), dispensing date, quantity, and defined daily doses. However, it does not include information on over-the-counter medications or drugs administered in hospitals. Together, these databases capture the entirety of government-funded healthcare utilization across both primary and specialist care sectors. ^21^

### Outcome

The primary outcome of this study will be spine-related healthcare costs (expressed in euros) incurred within 12 months following the index consultation. Healthcare utilization due to back and neck conditions will be classified based on the International Classification of Primary Care (ICPC-2) and the International Classification of Diseases (ICD-10) diagnostic medical codes (Appendix 1). Costs will be estimated using reimbursement rates from Norwegian Control and Payment of Health Reimbursements Database, the Norwegian Patient Registry, and the Norwegian Prescription Database, summarized per individual, and adjusted to 2022 price levels.

### Predictors

Table 2 reports the variables that will be considered as potential prognostic factors.

**TABLE 2.**
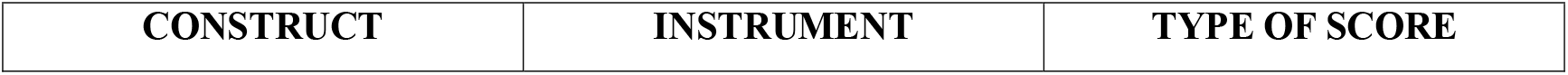
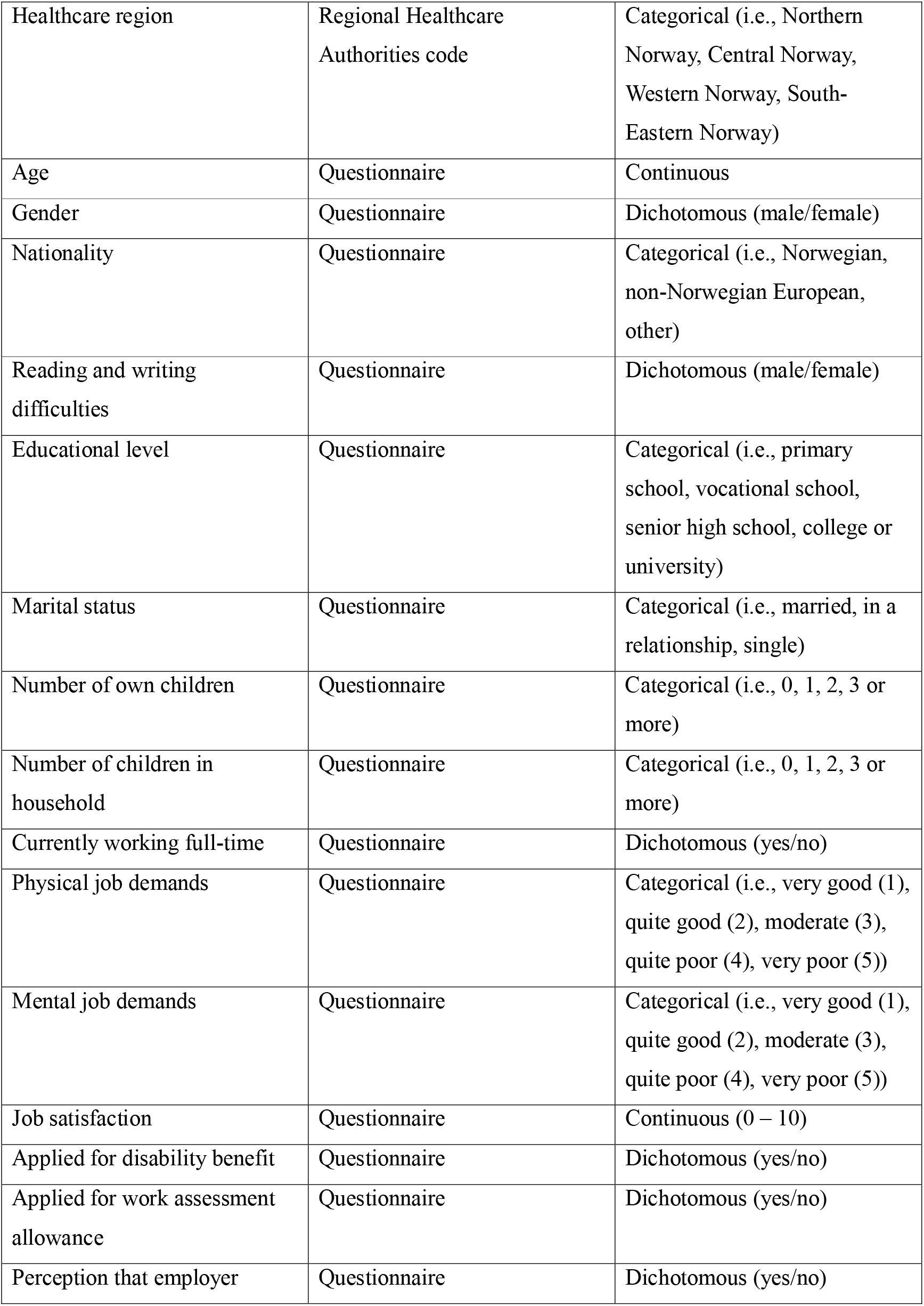
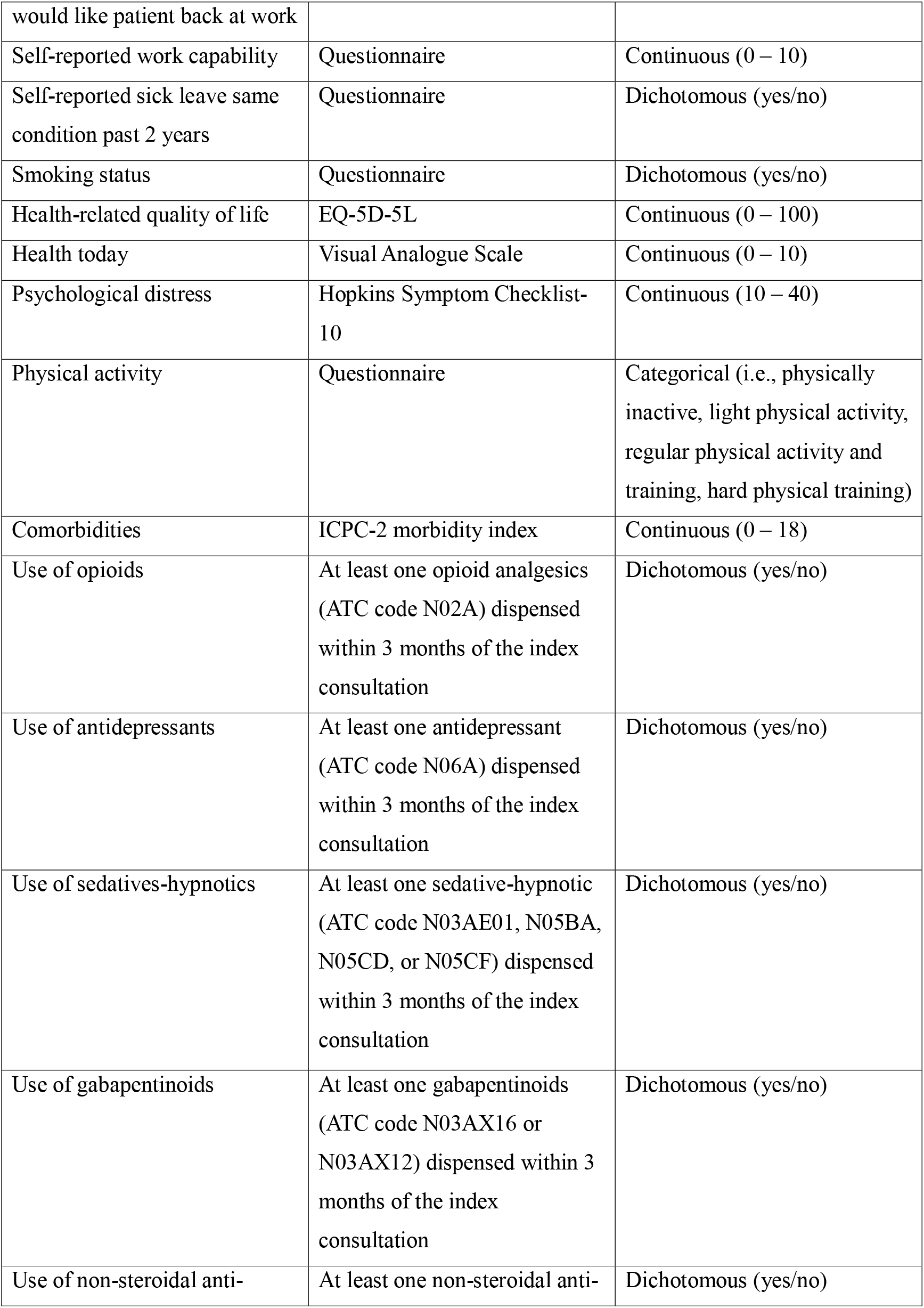
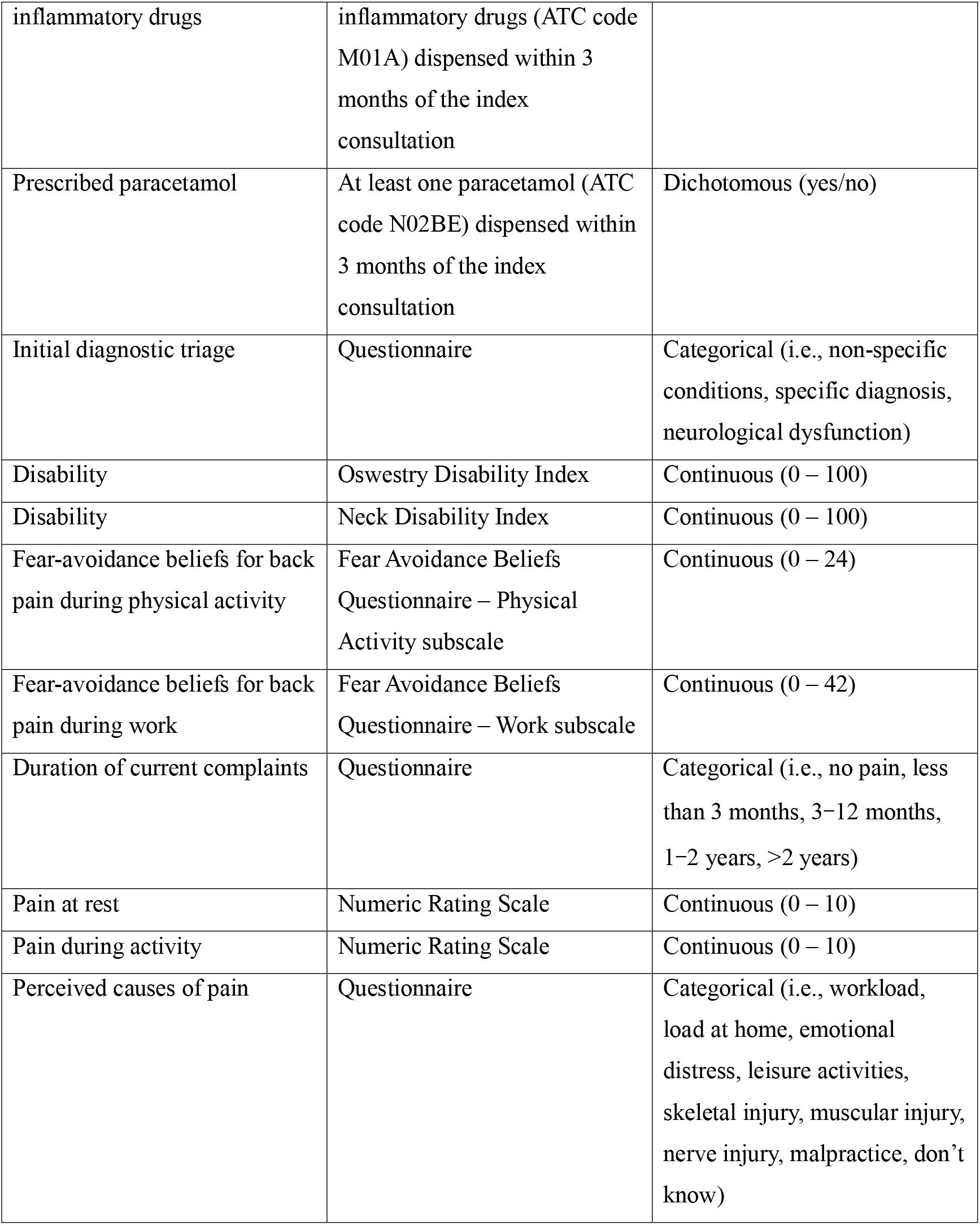
LIST OF CANDIDATE PREDICTORS.

### Statistical analysis

All statistical analyses will be conducted using R using the mice, rms, car, influence.ME, dplyr, tidyr, tibble, and ggplot2 packages. ^22^ Descriptive statistics will be used to summarize the study variables. Continuous variables will be reported as means and standard deviations or medians and interquartile ranges, depending on the distribution. Categorical variables will be presented as absolute frequencies and percentages.

Missing variables will be handled by multiple imputations by chained equations, using logistic regressions for binary variables, Bayesian polytomous regressions for categorical variables with more than two categories, and predictive mean matching for continuous variables. ^23^ We will use all the variables mentioned in Tables 1 to 5 for the imputation model, except for variables that have a high correlation (i.e., Spearman correlation coefficient > 0.9) or a high percentage of missing values (i.e., > 50%). ^23^ We will impute a number of datasets equal to the percentage of the most missing variable, using 20 iterations. ^23^ Convergence will be assessed visually, and the imputed data will be compared to the observed data to determine the plausibility of the imputed datasets. ^23^ The parameters of substantive interest will be estimated in each imputed dataset separately and combined using Rubin’s rules. ^23^

We anticipate that the outcome will be right-skewed. Therefore, the univariable associations between spinal healthcare utilization costs and each predictor will be assessed using a generalized linear model with a Gamma distribution. ^24^ For predictors that show a statistically significant association in the univariable analysis, we will conduct two additional adjusted analyses. The first will be adjusted for spinal healthcare utilization costs in the year preceding the index consultation, as well as for age, gender, baseline pain intensity, and baseline disability. These confounders were selected based on background knowledge and previous literature in the field. ^10,16,25–27^ The second analysis will include the same confounding variables, along with all other predictors that were statistically significant in the univariable analysis.

In all analyses, we will assume a smooth relationship of the continuous covariates using restricted cubic regression splines with three knots placed at the 10^th^, 50^th^, and 90^th^ percentile. ^28^ Categorical ordinal variables (i.e., educational level, physical activity, and duration of current complaints) will be converted into continuous variables and modelled accordingly with restricted cubic splines. ^28^ We will assess the assumption of the absence of multicollinearity using the Variance Inflation Factor with a cut-off of 5. In addition, we will evaluate the lack of influential observations using the DFBETAS with a cut-off of 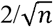. ^28^

For categorical variables with more than two categories and continuous variables (modelled using restricted cubic splines), overall (omnibus) Wald tests will be used to assess their statistical significance.

As subgroup analyses, we will explore outcomes separately based on pain localization (neck pain only, LBP only, and combined) as well as by specific categories of healthcare utilization. Also, as a sensitivity analysis, we will perform all multivariable models using a complete-case approach.

### Sample size

There are no closed-form formulas available for calculating the required sample size in multivariable prognostic factor studies. ^29^ Also, conducting a simulation-based sample size estimation is not feasible in this context, as it would require assumptions about the distribution of all candidate predictors and their inter-correlations—information that is currently unknown and difficult to approximate reliably. Therefore, we will adopt a commonly used rule of thumb, which suggests an effective sample size of at least 10 per every coefficient included in the final model. ^28^ In the case that all the candidate predictors are statistically significantly associated with the outcome, the final adjusted model will include a total of 66 coefficients in our model (40 from continuous variables and 25 from non-ordinal categorical variables). Accordingly, our sample size exceeds the minimum requirement of 660 patients to accurately estimate the model parameters based on the aforementioned rule of thumb.

## DISCUSSION

This study aims to identify prognostic factors associated with high healthcare utilization costs among patients referred to a specialist assessment/intervention in secondary care due to back and/or neck pain. The rationale stems from the well-established evidence that spinal pain conditions are highly prevalent and economically burdensome and that a small proportion of patients appears to account for the majority of healthcare costs, emphasizing the need to better understand which patient characteristics predict such disproportionate utilization. ^1,2,5,6,9,15^

A significant strength of this study is the use of the NNRR, a large, nationwide clinical registry that includes detailed, longitudinal, and systematically collected data from over 7,000 patients across each health region across Norway. ^20^ The registry provides a unique opportunity to explore a broad set of clinical, psychosocial, demographic, and work-related variables as potential prognostic factors. The analytical approach will be rigorous and transparent, adhering to the PROGRESS framework for prognostic research. ^17,18^ The use of multiple imputation, flexible modelling with restricted cubic splines, and sensitivity and subgroup analyses will further enhance the robustness of the study design. ^28^ Nonetheless, this study has some limitations. First, the generalizability of the findings may be restricted to healthcare systems with similar structures and registry infrastructures. Second, costs related to private healthcare utilization will not be included due to data unavailability. Third, costs associated with diagnostic imaging and rehabilitation services will be excluded, as these are incompletely reported in both the Norwegian Control and Payment of Health Reimbursements Database and the Norwegian Patient Registry during the study period. As a result, the total costs per patient may be underestimated.

## DECLARATION OF GENERATIVE AI AND AI-ASSISTED TECHNOLOGIES IN THE WRITING PROCESS

During the preparation of this work the author(s) used Grammarly and ChatGPT in order to check the syntax and grammar of the text. After using this tool/service, the author(s) reviewed and edited the content as needed and take(s) full responsibility for the content of the publication.

## Data Availability

All data produced in the present work are contained in the manuscript

---

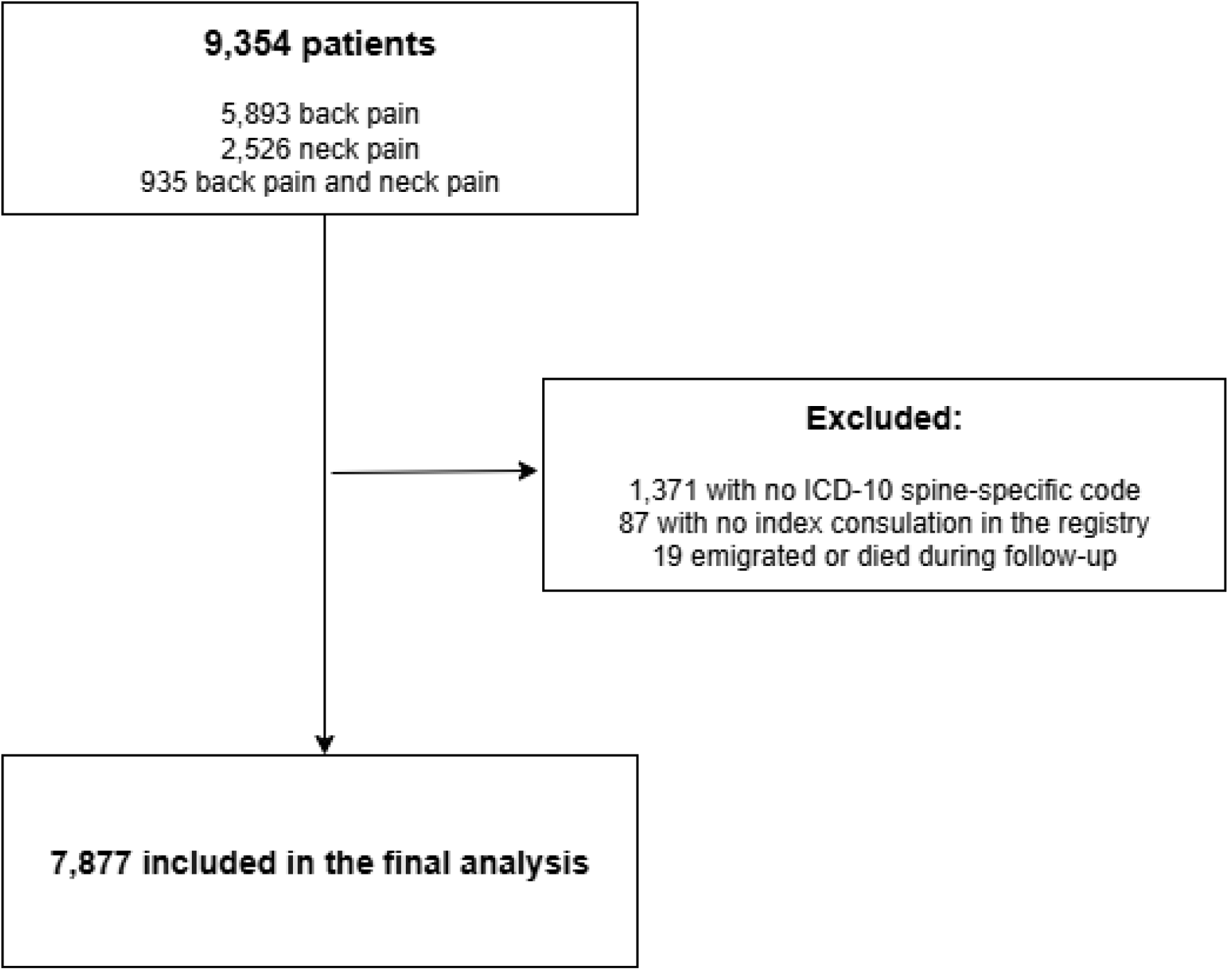

