## Appendix 1 for "Study Protocol: Prognostic Factors of High Healthcare Utilization Costs Among People with Spinal Disorders Using a Spine Registry Linked to Public Healthcare Data"

**APPENDIX 1 – ICD-10 CODES EXCLUDED FROM OUR ANALYSIS**

| **ICD-10 CODE** | **ICD-10 DESCRIPTION** | **FREQUENCY** |
| --- | --- | --- |
| M79.1 | Myalgia | 200 |
| M25.5 | Pain in joint | 19 |
| M76.0 | Gluteal tendinitis | 18 |
| M75.9 | Shoulder lesion, unspecified | 17 |
| M79.7 | Fibromyalgia | 15 |
| M45 | Ankylosing spondylitis | 15 |
| M75.1 | Rotator cuff syndrome | 13 |
| M75.4 | Impingement syndrome of the shoulder | 10 |
| S22.0 | Fracture of thoracic vertebra | 10 |
| M79.6 | Pain in limb | 9 |
| M70.6 | Throcanteric bursitis | 7 |
| M25.9 | Joint disorder, unspecified | 6 |
| S32.0 | Fracture of lumbar vertebra | 6 |
| G44.2 | Tension-type headache | 6 |
| M46.1 | Sacroiliitis, not elsewhere classified | 5 |
| S32 | Fracture of lumbar spine and pelvis | 5 |
| M75.0 | Adhesive capsulitis of shoulder | 5 |
| R51 | Headache | 4 |
| M46.8 | Other specified inflammatory spondylopathies | 4 |
| M16.9 | Coxarthrosis, unspecified | 3 |
| R42 | Dizziness and giddiness | 3 |
| R52.9 | Pain, unspecified | 3 |
| T91.1 | Sequelae of fracture of spine | 3 |
| M75.8 | Other shoulder lesions | 3 |
| M41.1 | Juvenile idiopathic scoliosis | 2 |
| M16.1 | Other primary coxarthrosis | 2 |
| S33.1 | Dislocation of lumbar vertebra | 2 |
| M54 | Dorsalgia | 2 |
| M62.9 | Disorder of muscle, unspecified | 2 |
| M46.0 | Spinal enthesopathy | 2 |
| M77.1 | Lateral epicondylitis | 2 |
| M22.2 | Patellofemoral disorders | 1 |
| G62.9 | Polyneuropathy, unspecified | 1 |
| L40.5 | Arthropathic psoriasis | 1 |
| R26.2 | Difficulty in walking, not elsewhere classified | 1 |
| M79.8 | Other specified soft tissue disorders | 1 |
| M15.9 | Polyarthrosis, unspecified | 1 |
| G54.0 | Brachial plexus disorders | 1 |
| M16.2 | Coxarthrosis resulting from dysplasia, bilateral | 1 |
| R52.2 | Other chronic pain | 1 |
| M18.9 | Arthrosis of first carpometacarpal joint, unspecified | 1 |
| G56.8 | Other mononeuropathies of upper limb | 1 |
| M94.0 | Chondrocostal junction syndrome | 1 |
| M70.9 | Unspecified soft tissue disorder related to use, overuse and pressure | 1 |
| M25 | Other joint disorder, not elsewhere classified | 1 |
| M46.4 | Discitis, unspecified | 1 |
| S39.0 | Injury of muscle and tendon of abdomen, lower back and pelvis | 1 |
| S12.2 | Fracture of other specified cervical vertebra | 1 |
| G57.1 | Meralgia paraesthetica | 1 |
| M48.5 | Collapsed vertebra, not elsewhere classified | 1 |
| F45.8 | Other somatoform disorders | 1 |
| S32.8 | Fracture of other and unspecified parts of lumbar spine and pelvis | 1 |
| G35 | Multiple sclerosis | 1 |
| G56.0 | Carpal tunnel syndrome | 1 |
| G43.1 | Migraine with aura | 1 |
| M54.0 | Panniculitis affecting regions of the neck and back | 1 |
| R52.1 | Chronic intractable pain | 1 |
| M42.1 | Adult osteochonrosis of spine | 1 |
| M65.9 | Synovitis and tenosynovitis, unspecified | 1 |
| S13.4 | Sprain and strain of cervical spine | 1 |
| M42.0 | Juvenile osteochonrosis of spine | 1 |
| M46.9 | Inflammatory spondylopathy, unspecified | 1 |
| F48 | Neurasthenia | 1 |
| S22.1 | Multiple fractures of thoracic spine | 1 |
| G43.0 | Migraine without aura | 1 |
| M19.0 | Primary arthrosis of other joints | 1 |
| M70.7 | Other bursitis of hip | 1 |
| M41.3 | Thoracogenic scoliosis | 1 |
